# Reduced reliance on partner reciprocity drives lower trust in schizophrenia

**DOI:** 10.1101/2025.11.06.25339661

**Authors:** Gabriele Bellucci, Lucy Vanes, Francesco Scaramozzino, Nicholas Furl, Sukhi S. Shergill

**Affiliations:** Center for Decision Sciences, Royal Holloway, University of London, Egham TW20 0EX; Department of Psychology, Royal Holloway, University of London, Egham TW20 0EX, UK; Department of Neuroimaging, Institute of Psychiatry, Psychology & Neuroscience, King’s College London, London, UK; Kent and Medway Medical School, University of Kent, Canterbury, UK; Department of Psychosis Studies, Institute of Psychiatry, Psychology & Neuroscience, King’s College London, London, UK

## Abstract

Schizophrenia is characterized by significant cognitive and social difficulties, including paranoid ideation and impaired trust in others. Despite consistent evidence of disrupted social interactions in schizophrenia, the neurocognitive mechanisms driving these impairments remain poorly understood. Here, we tested the hypothesis that schizophrenia is associated with altered neural processing of others’ cooperative behavior, leading to disrupted belief formation and reduced trust. In a sample of individuals with a diagnosis of schizophrenia (*N*=39) and healthy controls (*N*=39) playing a computerized investment game, we show that individuals affected by schizophrenia trusted their game partners less, which was due to reduced sensitivity to partner reciprocity and greater variability in trust behavior. Computational modeling shows that, while both groups learned the partner’s behavior equally well, individuals with schizophrenia were less likely to act on their beliefs about partner benevolence. Neuroimaging results show that during encoding of partner reciprocity, the putamen was more engaged and functionally connected with the precuneus in healthy controls. Reduced functional connectivity between putamen and precuneus was further associated with less successful trusting interactions (lower payoffs) in individuals with schizophrenia as compared with control participants. These results indicate how belief formation impact social behaviors such as trust in clinical populations.

## Introduction

Trust is defined as the willingness to accept vulnerability based on one’s expectations of others’ intentions and behaviors^1–3^. Hence, decisions to trust depend on an individual’s perceptions of the vulnerable state that follows trust, and on an individual’s perceptions of the others’ trustworthiness. Clearly, expecting someone to behave unreliably or to be ill-intentioned deters trust, as the potential negative consequences of becoming dependent on that person weigh in favor of withdrawing from the interaction. Importantly, however, cases of misplaced distrust have been understudied in the literature. In such cases, the other person’s trustworthiness (e.g., their benevolence) would suggest that trust is warranted, yet individuals still choose to distrust them. These instances highlight that trust is not merely the outcome of an economic cost-benefit analysis based on one’s beliefs about a partner’s trustworthiness but also depends on individual preferences in interpersonal relationships.

Distrusting ostensibly trustworthy others may significantly hinder an individual’s ability to form social connections and be a behavioral marker of potential mental health issues^4^. For instance, schizophrenia (SZ) is characterized by a complex interplay of positive (hallucinations and delusions) and negative (affective disengagement and social withdrawal) symptoms that profoundly affect social and occupational functioning, including trust^5^. Social withdrawal and paranoid thoughts are core, debilitating symptoms of the disorder^8,9^. Negative expectations and negative evaluations of others may undermine social motivation and contribute to social exclusion and broader difficulties in social functioning^1–3^. This, in turn, exacerbates psychotic experiences and paranoid delusions^6–8^. Individuals with SZ have been reported to experience frequent paranoid thoughts, which foster social withdrawal and isolation^9,10^ and prior research has shown that individuals with psychosis exhibit issues with trust^10–13^.

These findings align with initial evidence on the mechanisms underlying the development of interpersonal trust in individuals with SZ. For instance, prior work has shown that individuals with SZ demonstrate lower levels of cooperation and fail to accurately adjust their behavior in response to social feedback^11,12^. It has been suggested that paranoid thoughts may contribute to such behavioral rigidity and maladaptive belief updating. Supporting this suggestion, paranoia has been associated with a stronger adherence to priors about environmental volatility in non-social tasks, leading to impaired belief updating and heightened sensitivity to perceived environmental changes^13^. Moreover, paranoid ideation has been linked to stronger prior beliefs about others’ harmful intent and greater policy uncertainty in social economic tasks^14^. These findings suggest that in trust-based interactions, individuals with paranoia may hold stronger priors about others’ untrustworthiness and exhibit heightened sensitivity to changes in cooperative behavior—particularly to reductions in trustworthiness. However, although a fundamental lack of trust has long been clinically viewed as a key factor underlying psychotic symptoms like paranoid delusions^15^, cognitive models of SZ have largely overlooked trust^16^.

In healthy individuals, trust has been associated with specific brain regions that show altered functioning in individuals with SZ. Neuroimaging studies, for example, have identified areas such as the medial prefrontal cortex (MPFC) and the precuneus—both part of the default mode network—as being related to social cognition and predictive of trusting behaviors^17^. Studies using the investment game to examine reciprocal trust have found that the anterior insula (AI) is consistently activated when individuals trust strangers in single interactions^18^, likely reflecting a heightened fear of betrayal^19^. By contrast, the dorsal striatum (i.e., caudate and putamen) has been associated with learning processes during repeated interactions^20^ and has been shown to track the learned reciprocity of a partner^21^.

Altered neural responses in the dopaminergic system including the striatum have long been implicated in SZ^22^. In non-social tasks, individuals with SZ demonstrate weaker recruitment of the striatum during reward anticipation and reward learning, with this attenuated response correlating with the severity of positive symptoms^23,24^. Furthermore, dopamine dysfunction has not only been linked to altered learning but also to social processing in SZ. For example, individuals with SZ show reduced caudate activation in response to partner cooperation, suggesting reduced social reward sensitivity^25^. Similarly, abnormal activity in the AI and precuneus have also been associated with social cognitive deficits^26^. In particular, reduced activation in the precuneus has been linked to diminished empathic accuracy in individuals with SZ^27^. However, what remains poorly understood is how these altered neural responses give rise to maladaptive behavioral patterns such as distrust in trustworthy others. In particular, the cognitive and computational mechanisms that link brain function to distrust-related behavior, such as how expectations about others are encoded, updated, and acted upon in SZ, are still unclear. Fine-grained computational approaches offer a promising means of formalizing these processes and identifying how disruptions in specific parameters map on to both brain activity and behavior. In this study, we combined the investment game (**Fig. 1A-B**) with neuroimaging and Bayesian computational modeling to investigate whether distrust in individuals with SZ (*N* = 39) is due to differences in how they weigh expectations of others’ trustworthiness when making trust decisions as compared to healthy controls (*N* = 39). All participants played as investors, interacting with a trustee who was preprogrammed to reciprocate benevolently. This setup allowed us to control for trustee trustworthiness and attribute any differences in trust behavior to participants’ learning and inference processes. Participants’ investments are classically taken as a measure of their trust, while trustees’ repayments as a measure of reciprocity. Computational modeling enabled us to formalize participants’ beliefs about the trustee and estimate their trial-by-trial decision policies, shedding light on their trust strategies during the task. We hypothesized that processing of others’ behavior in brain regions associated with reward processing and social cognition would be altered in individuals with SZ, preventing them from accurately learning others’ trustworthiness for successful trusting interactions.

**Fig. 1.**
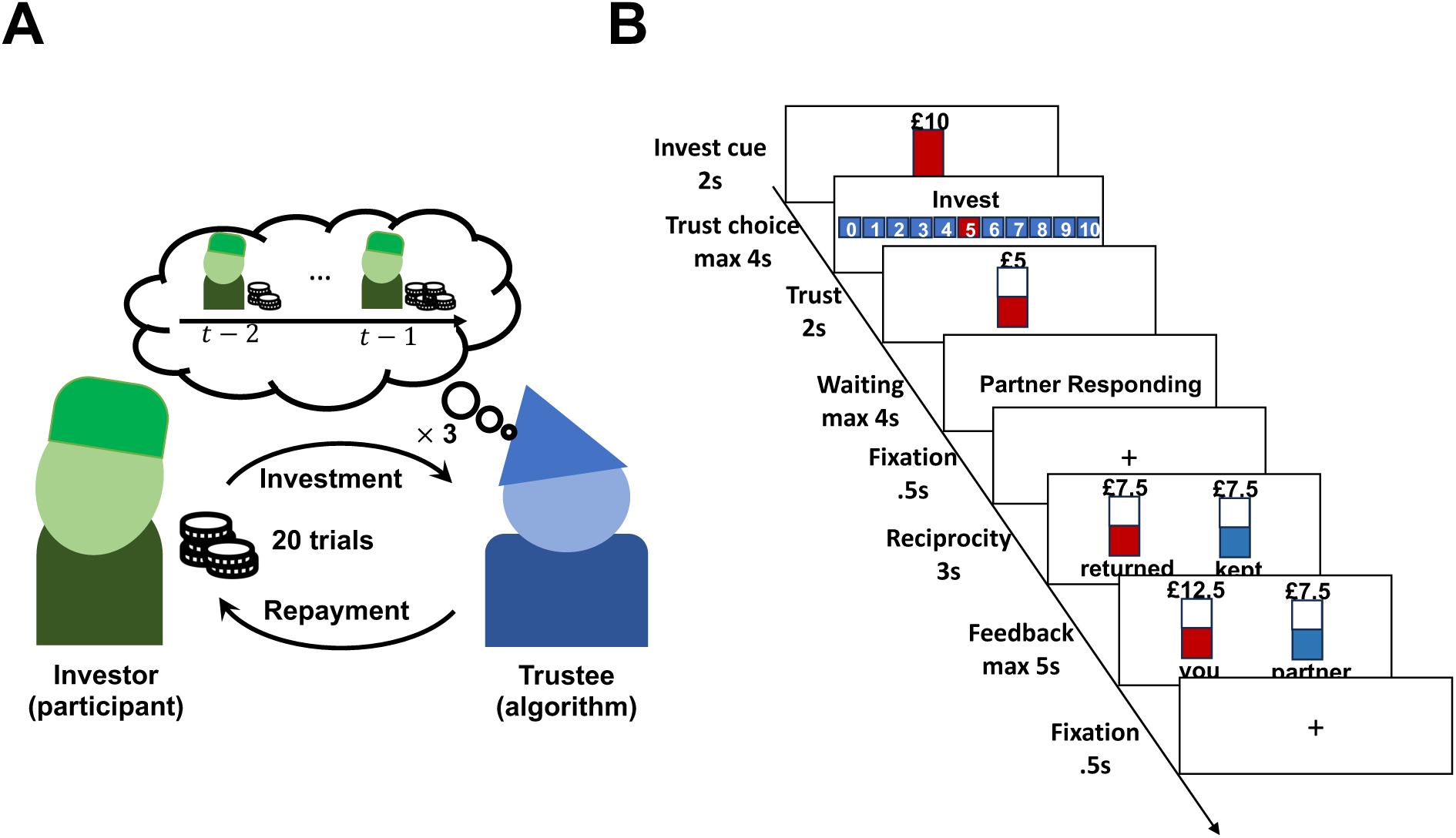
Task and experimental design. **A.** Participants played 20 trials of the investment game with the same trustee. Trustee behavior was simulated by an algorithm that was preprogrammed to play benevolently. Investments by the participant are considered as a measure of trust, repayments by the trustee as a measure of reciprocity. **B.** Experimental design in the MRI scanner: at the beginning of every trial (investment cue), participants received an initial endowment (£10) and have to choose whether to share any of this amount with the trustee (trust choice). The investment is shown on the screen (trust) and is tripled before being passed on to the trustee. The participant waits until the trustee decides whether to repay any amount (waiting). After a short fixation, the repayment is shown (reciprocity) before the amount earned by both players is presented (feedback).

## Results

Each participant played the investment game as an investor for 20 trials. The trustees were simulated by a computer programmed to behave in a probabilistically benevolent manner (**Fig. 1A**). The trustees simulated the behavior of a benevolent partner by rewarding increases of trust with greater reciprocity, while decreases of trust had no negative impact on reciprocity (see *Methods*).

### Lower trust in individuals with SZ

Individuals with SZ exhibited lower average levels of trust across the task than controls (*t*_(76)_ = −2.61, *p* = .011; **Fig. 2A**). In turn, these lower levels of trust by our participants induced lower levels of reciprocity by trustees (*t*_(76)_= −2.60, *p* = .0114; **Fig. 2B**). Importantly, while first trust choices on the first trial were comparable between groups (*t*_(76)_= −1.22, *p* = .229), trust in the last trials was significantly greater for controls than for individuals with SZ (*t*_(76)_ = −3.60, *p* < .0006). A mixed-effects Bayesian regression to predict trialwise trust using previous reciprocity (*rec*_*t*−1_), group and time as predictors shows that trust increased over time (β_*time*_ = 0.14, 95% highest density intervals, HDI, = [0.06, 0.22]) but less so for SZ patients (β_*group×time*_ = −0.12, 95% HDI = [−0.23, −0.01]; **Fig. 2C**). Specifically, while participants on average responded to greater reciprocity with greater trust (β_*rec×t*−1_ = 0.18, 95% HDI = [0.10, 0.26]), SZ patients were less responsive to trustee reciprocity (β_*group×rec×t*−1_ = −0.13, 95% HDI = [−0.24, −0.02]). Further, sensitivity to trustee reciprocity increased over time more strongly in controls than SZ patients (β_*group×time*×*rec×t*−1_ = 0.01, 95% HDI = [0.00, 0.02]; **Fig. 2D**), indicating successful learning about trustee reciprocity in controls over time. Finally, SZ patients’ trust manifested higher variability (*t*_(76)_ = 2.15, *p* = .035; **Fig. 2D**) and showed greater change across trials (*t*_(76)_ = 2.54, *p* = .013; **Fig. 2E**), indicating less stable and more uncertain trusting behaviors.

**Fig. 2.**
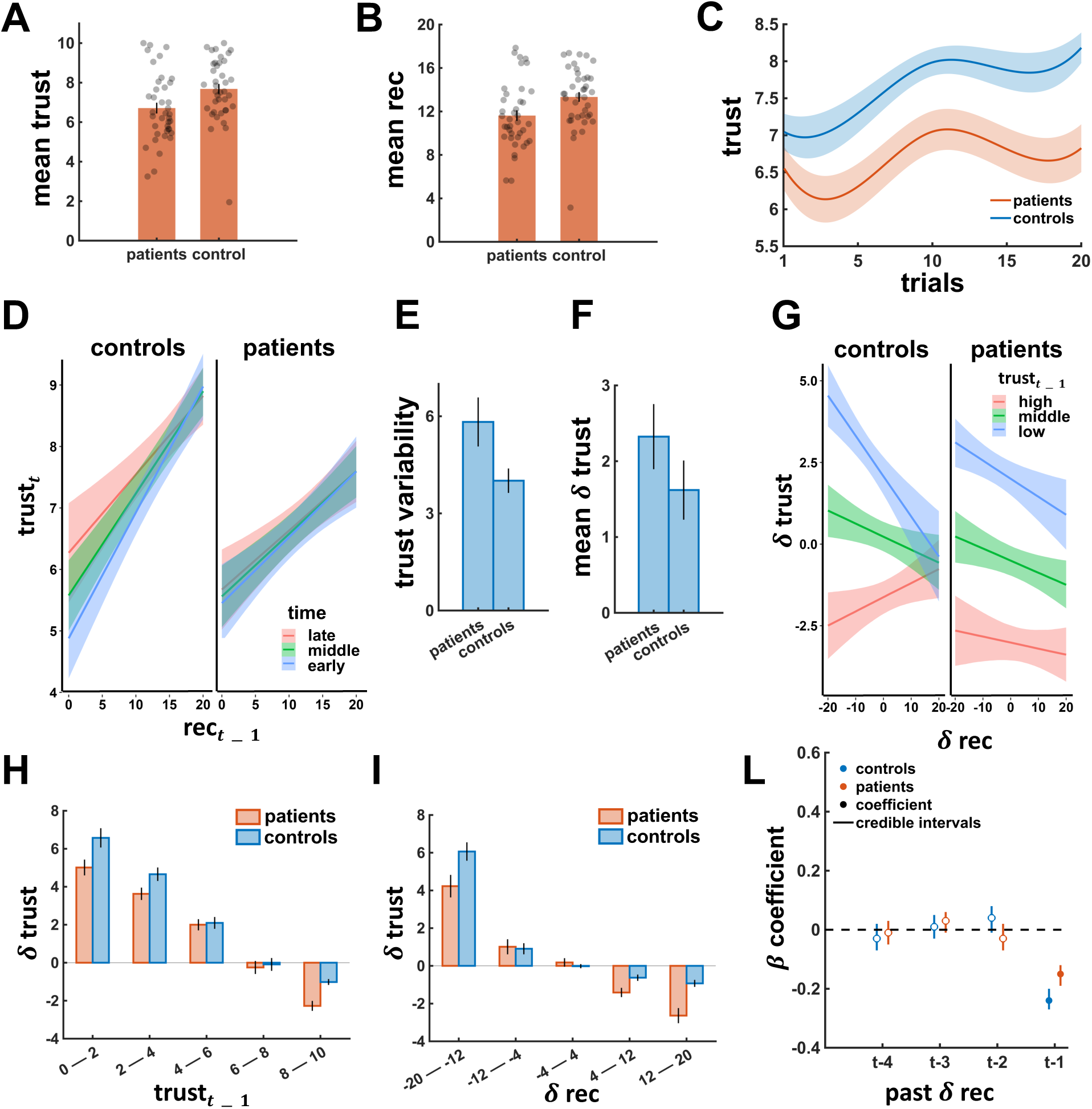
Trust in individuals with SZ and controls. **A.** Individuals with SZ had lower levels of average trust than controls. **B.** Lower levels of trust in individuals with SZ led the algorithm to reciprocate less as compared to interactions with controls. **C.** Over time, controls but not individuals with SZ increased their trust. Shaded areas represent standard error of the mean. **D.** Compared with individuals with SZ, controls trusted lowly-reciprocating trustees more toward the end of the investment game (i.e., late). Shaded areas are 95% HDIs. **E.** Trusting behavior in individuals with SZ had a higher variability than controls’. **F.** Average trust changes (δ_*t*_ = *trust*_*t*_ − *trust*_*t*−1_) were higher in individuals with SZ than controls. **G.** Controls were more sensitive to changes of partner reciprocity, sharing more in response to positive reciprocity, while individuals with SZ showed overall distrustful behavior. Shaded areas are 95% HDIs **H.** Lower levels of trust in the previous trial (*trust*_*t*−1_) predicted stronger increases of future trust in controls than individuals with SZ, while higher levels of trust predicted stronger decreases of future trust in individuals with SZ than controls. **I.** Controls showed stronger coaxing behaviors (increases of trust in response to negative reciprocity) than individuals with SZ. **L.** In both individuals with SZ and controls, trust changes were best predicted by the most recent changes of partner reciprocity, suggesting shorter behavioral dependencies (parameter values are regression coefficient values). rec, reciprocity; HDI, highest density intervals. Error bars represent standard error of the mean.

### Lower sensitivity to trustee trustworthiness in individuals with SZ

Even though our preprogrammed trustees behaved similarly across groups (*t*_(76)_= −1.35, *p* = .182), we observed that individuals with SZ changed their trust more strongly than controls in response to trustee reciprocity changes (β_*group×rec*_ = 0.08, 95% HDI = [0.06, 0.11]), leading to greater trust changes on average (**Fig. 2F**). We hence investigated conditional trust changes as a model-agnostic measure of an individual’s sensitivity to trustee trustworthiness with a mixed-effects Bayesian regression to predict the last two-trial trust change (δ_*trust*_) using group, previous trust (i.e., trust on the previous trial: *trust*_*t*−1_) and the last two-trial reciprocity change (δ_*rec*_) as predictors.

We found that stronger increases of trust were overall triggered by sharper decreases of reciprocity, likely to “coax” the trustee to cooperate (β_δrec_ = −0.26, 95% HDI = [−0.36, −0.16]). We also found that higher levels of previous trust predicted smaller increases of trust, likely due to ceiling effects induced by the learning dynamics (β_*trust×t*−1_ = −0.67, 95% HDI = [−0.78, −0.56]). Moreover, trust changes in response to reciprocity changes differed across groups as a function of previous trust (β_*group*δrec×*trust*×t−1_ = −0.02, 95% HDI = [−0.04, −0.01]; **Fig. 2G**). Specifically, controls who shared more on the previous trial were more likely to increase their trust in response to positive reciprocity (positive δ_*rec*_; **Fig. 2H-I**), indicating higher responsiveness to trustees’ signals of trustworthiness. In contrast, individuals with SZ were less sensitive to trustee reciprocity, less robustly coaxed cooperation, and were more likely to reduce high levels of trust in response to positive reciprocity (**Fig. 2H-I**).

Finally, trust changes in response to reciprocity changes revealed recent-history dependencies (**Fig. 2L**). The effect decreased from β = −0.26 to β = −0.02 when considering reciprocity changes further back in time. Moreover, only the last two-trial reciprocity change (δ_*rec×t*−1_) significantly predicted the last two-trial trust change in both controls (β = −0.24, 95% HDI = [−0.27, −0.20]) and individuals with SZ (β = −0.15, 95% HDI = [−0.19, −0.12]) with individuals with SZ showing a smaller effect size in line with their lower sensitivity to trustee trustworthiness. In line with previous work^20,28^, these findings indicate that investors’ trust in the investment game is most strongly influenced by the trustee’s most recent behavioral choices.

### Computational modeling

We investigated these sequential effects by formalizing how participants learned about their partners over time and how much weight they put on trustee behavior. We considered two main classes of model and their variations (see *Methods*; **Tab. S1**). The first was the vulnerability model, which has been proposed to closely capture psychological and computational components of trust (Fig. 3A)^3^. Following classic definitions of trust^1,2^, the vulnerability model defines trust as a tradeoff between the potential negative consequences of trusting (in the investment game, the possibility of losing the investment *i*_*t*_) and the expected positive intentions of others (in the investment game, the trustee’s repayment *r*_*t*_). A participant-specific parameter (τ ∈ [0,1]) captures an individual’s willingness to rely on one’s expectations of the other’s intentions at the cost of making oneself vulnerable to them (i.e., willingness to trust). In our study, participants’ expectations are represented as a probability distribution over all possible repayments contingent on participants’ current investment and the history of interactions with the trustee, that is, *p*(*R*_*t*_ = *r*_*t*_|*D*_*t*−1_), where *D*_*t*−1_ includes the history of repayments until time *t* (i.e., *r*_1:*t*−1_). As proportions naturally range between 0 and 1 so that *r*_*t*_ ∈ [0,1], *p*(*R*_*t*_ = *r*_*t*_|*D*_*t*_) takes the form of a beta distribution. Action utilities can be described with the following utility function:

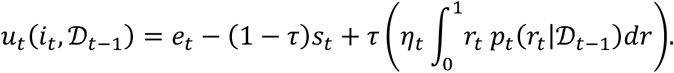

**Figure 3.**
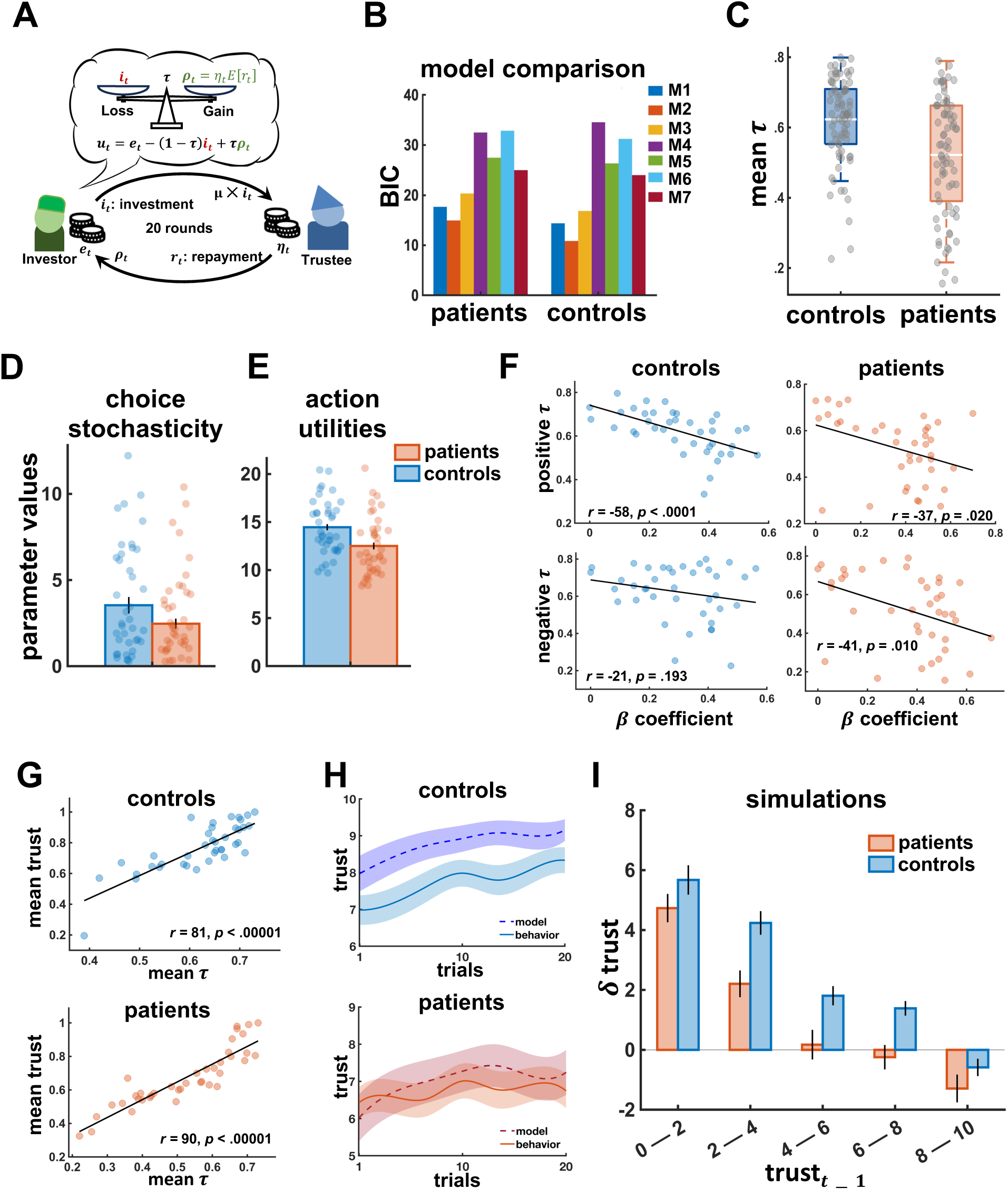
Modeling results. **A.** The winning model was one that weights the potential loss of trusting (the amount shared) against the expected benefits of reciprocity (the expected repayment from the trustee). The weighting (i.e., τ) parameter is taken to capture individual differences in participants’ willingness to trust. **B.** Model comparison shows that the best model in both controls and individuals with SZ was the “vulnerability model” weighting expectations of partner reciprocity differently after positive and negative reciprocity (i.e., M2). **C.** Controls were overall more willing to trust their trustees than patients (had higher τ parameter values). **D.** We observed a trend toward lower choice stochasticity (i.e., χ parameter in the model’s policy) in individuals with SZ than controls, indicating greater decision noise in individuals with SZ. **E.** Trialwise action values were on average lower for individuals with SZ than controls. **F.** The magnitude of trust changes (β coefficients) negatively correlated with individuals with SZ willingness to trust both after positive (positive τ) and negative (negative τ) reciprocity, suggesting that less trusting patients changed their trust to a greater degree trial-by-trial, while controls showed this effect only after positive reciprocity. **G.** Average willingness to trust after positive and negative reciprocity (mean τ) predicted higher average trust levels (investments). **H.** Simulated behavior based on τ parameters fitted to controls manifested the empirical increase of trust observed over time in controls, while simulated behavior based on τ parameters fitted to individuals with SZ shows similar trust levels across the whole game as empirically observed in individuals with SZ. Shaded areas represent standard errors. **I.** Simulated behavior replicated the interaction effect between group and previous trust levels as observed in the empirical data in Fig. 2H. BIC, Bayesian Information Criterion. Error bars represent standard error of the mean.

Prior expectations (ρ_0_) before the interaction (i.e., *D*_*t*_ = ∅, for *t* = 1) were estimated from the data and updated trial by trial following Bayes’ rule. Choices were assumed to be made stochastically based on participants’ exponentiated action utilities, so that action probabilities follow a softmax function:

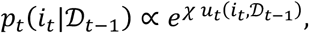

where χ is a participant-specific parameter that captures their choice consistency (stochasticity). The second class of models was the inequity-aversion model^29^, which captures individual aversion to unequal outcomes in resource distributions (see *Methods*)^30^. We also considered a simpler Bayesian imitator model for model comparisons (**Tab. S1**). The winning model was a vulnerability model with two weighting parameters for participants’ expectations after positive and negative reciprocity (M2 in Fig. 3B), capturing the sequential effects observed in the behavioral analyses. This model’s root mean squared error was 2.9 and recovery analyses indicated high model and parameter recoverability (*Methods*).

### Lower willingness to trust in individuals with SZ

Prior expectations (ρ_0_) strongly correlated with participants’ first choice in the first trial in both groups (controls: *r_37_* = .71, *p* < .00001; patients: *r_37_* = .58, *p* < .00001) but did not significantly differ across groups (*t*_(76)_ = −0.09, *p* = .930), suggesting that the model effectively captured participants’ expectations of others’ trustworthiness relevant for trust. Indeed, individuals with SZ (ρ_0_ = .44, SD = .15) and controls (ρ_0_ = .45, SD = .17) both expected their partner to be fairly trustworthy at the beginning of the interaction. However, a participant-level Bayesian regression to predict individual willingness to trust (i.e., τ) using group and τ type (i.e., τ_*p*_and τ_*n*_) revealed that the groups significantly differed in their willingness to trust with individuals with SZ underweighting their expectations of trustee reciprocity more strongly than controls (β_*group*_= −0.08, 95% HDI = [−0.16, −0.01]; Fig. 3C), even though both groups’ willingness to trust was comparable after positive (τ_*p*_) and negative (τ_*n*_) reciprocity (β_*group*τ_= 0.004, 95% HDI = [−0.08, 0.08]).

Further, the above-mentioned higher trust variability in individuals with SZ (Fig. 2E) could have meant nosier choice patterns (constantly changing the amounts shared) or more deterministic ones (switching between extremes). Analyses of the stochasticity parameter (i.e., χ) suggest the former hypothesis, indicating a trend in individuals with SZ towards lower choice consistency than controls (*t*_(70)_ = −1.84, *p* = .070; Fig. 3D). Moreover, model-based action utilities for the empirical choice at time *t* were consistently lower (*t*_(76)_= −2.94, *p* = .004; Fig. 3E) in individuals with SZ than controls. This finding indicates a lower contribution of expectations of trustee reciprocity to current trust decisions, whose values were more greatly influenced by the possibility of losing the investment *i*_*t*_ (i.e., the negative consequence of trusting). Moreover, action utilities across the action space had lower variance in individuals with SZ as compared to controls (*t*_(76)_ = −2.32, *p* = .023), indicating reduced expected-value differences between the different investment options in individuals with SZ.

Importantly, though, trust can be successfully established only if one adequately adapts to another’s trustworthiness and since in our task, trustees were preprogrammed to behave benevolently, it was beneficial to adapt to increases of trustee reciprocity. To examine participants’ ability to adequately adapt their trust to trustee reciprocity, we analyzed individual policy changes as estimated by the model (i.e., changes in action probability distributions). We found significant policy changes in controls after positive reciprocity (*t*_(3=)_= −2.42, *p* = .020), but not in SZ patients (*t*_(3=)_ = −0.48, *p* = .637). However, significant policy changes were found in both controls (*t*_(3=)_= 3.27, *p* = .002) and individuals with SZ (*t*_(3=)_= 2.74, *p* = .009) after negative reciprocity. Importantly, policy changes were positive for both controls (*M* = .07, SD =.13) and patients (*M* = .06, SD =.13) after negative reciprocity, suggesting that a previous trust choice was more likely to be repeated after decreases of reciprocity. On the contrary, policy changes were negative in controls (*M* = −.03, SD =.08) after positive reciprocity, indicating a higher likelihood of behavior change after increases of reciprocity. As policy changes were further positively correlated with individual trust changes in both controls (*r_76_* = .49, *p* < .00001) and individuals with SZ (*r_76_* = .34, *p* = .002), these findings suggest that both individuals with SZ and controls were more cautious after signs of decreasing reciprocity, but controls were also better able to quickly adapt their level of trust with partners who signaled a readiness to reciprocate.

Further, the magnitude of trust changes in response to reciprocity changes negatively correlated with participants’ willingness to trust (Fig. 3F). In individuals with SZ, that was the case both after positive (τ_*p*_: *r_37_* = −.37, *p* = .020) and negative (τ_*n*_: *r_37_* = −.41, *p* = .010) reciprocity, while in controls only after positive (*r_37_* = −.58, *p* < .0001) but not negative (*r_37_* = −.21, *p* < .193) reciprocity, suggesting that distrustful participants were more likely to probe their trustees with higher levels of trust. Further, lower average willingness to trust correlated with lower choice stochasticity values in both controls (*r_37_* = .34, *p* = .035) and individuals with SZ (*r_37_* = .40, *p* = .012), indicating noisier choice patterns in more distrustful participants.

This implies that participants with a lower willingness to trust exhibited overall lower levels of trust. This was indeed confirmed by a strong linear relationship between individual willingness to trust and average trust in both controls (*r_37_* = .81, *p* < .00001) and individuals with SZ (*r_37_* = .90, *p* < .00001; Fig. 3G). Moreover, the steady increase of trust over time in our controls could have depended on controls’ higher willingness to trust. Indeed, simulated behavior using participants’ fitted parameter estimates from the winning model shows that higher τ parameters in controls predict a steady increase of trust over time (higher panel in Fig. 3H). On the contrary, simulated behavior using τ parameters fitted to individuals with SZ predicted similar trust levels across the whole interaction (lower panel in Fig. 3H). This is because underweighting beliefs about trustee reciprocity (lower τ values) implied that trustee benevolence had a smaller impact on action utilities for higher levels of trust in individuals with SZ. In other words, it was hard for our distrustful individuals with SZ to trust their trustees––no matter how benevolently they behaved. Finally, Fig. 3H shows a good correspondence between true and simulated behavior, demonstrating that the model was able to faithfully capture other key behavioral effects of our empirical data, such as the correspondence between model-based expectations of trustee reciprocity and true reciprocity (controls: *r_37_* = .74, *p* < .003; patients: *r_37_* = .63, *p* < .003) and a significant difference in trust changes as a function of previous levels of trust between groups (β_*group×trSDt×t*−1_ = −0.10, 95% HDI = [−0.19, −0.01]; Fig. 3I).

### Different trust and feedback processing in SZ patients

Contrast analyses of neuroimaging data during trust choices (Fig. 1B) revealed that controls activated the caudate (extending to putamen) more strongly than patients (Fig. 4A), while the AI was more strongly recruited by individuals with SZ than controls after trust choices (Fig. 4B). Based on previous research^18^, we further tested whether neural activity shifts from AI to the striatum as an individual learns trustee trustworthiness over time and is able to predict beneficial consequences of trust. We observed that individuals with SZ showed stronger AI engagement compared to controls only in early trials, which then significantly decreased over time for individuals with SZ (β = −0.32, 95% HDI = [−0.59, −0.03]) but remained low and flat for controls (β = −0.08, 89% CI = [−0.27, 0.12]; lower panel of Fig. 4B).

**Figure 4.**
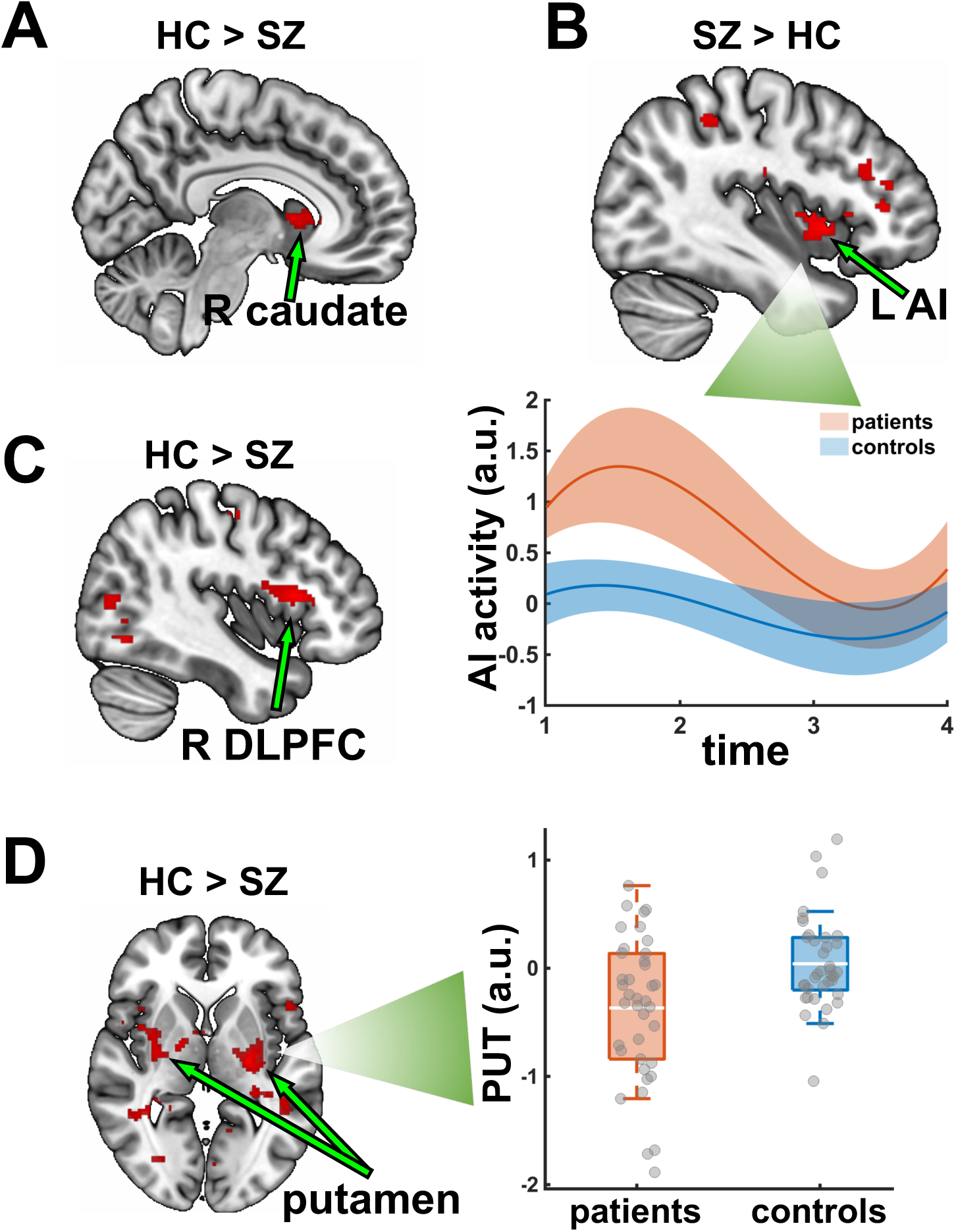
Group differences in neural activations. **A.** At trust choice, the right caudate/putamen (MNI: x = 16, y = 18, z = 2) was activated more in controls than patients. **B.** At trust, the left AI (MNI: x = −38, y = 10, z = −2) was activated more in individuals with SZ than controls, especially in the early trials (lower panel). **C.** At reciprocity, expectation violations activated the right DLPFC (MNI: x = 42, y = 18, z = 12) more in controls than patients. **D.** Payoff parametric modulation of feedback activity showed stronger engagement of the right (MNI: x = 32, y = −8, z = 0) and left (MNI: x = −28, y = −18, z = 4) putamen in controls than SZ patients. AI, anterior insula; DLPFC, dorsolateral prefrontal cortex; PUT, putamen; L, left; R, right; u.a., arbitrary units; shaded areas are standard errors. FWE<.05 cluster-level corrected.

Further, failures to appropriately adapt one’s trust to trustee reciprocity could be traced back to differential processing of feedback about trustee reciprocity. In particular, the above-mentioned lower sensitivity to trustee reciprocity in individuals with SZ (Fig. 2G) could have hinged on reduced neural responses to unexpected changes in partner behavior. Indeed, expectation violations (i.e., the difference between ρ_*t*_and *r*_*t*_, see Fig. 3A) during the reciprocity phase more strongly activated the right dorsolateral prefrontal cortex (DLPFC) and left precentral gyrus (MNI: x = −46, y = −16, z = 56) in controls than individuals with SZ (Fig. 4C). Finally, brain activity in the bilateral putamen during feedback more strongly correlated with one’s own payoffs in controls than individuals with SZ (Fig. 4D), suggesting group differences in reward processing that could have impacted trust learning and building over time.

### Relationships between willingness to trust and brain activity

Next, we investigated whether and how these brain regions were associated with individual willingness to trust by running whole-brain regression analyses with individual τ parameters as participant-level regressors. Group results showed that midcingulate cortex (MCC) at the beginning of the trial (Fig. 5A), left DLPFC and ventromedial prefrontal cortex (VMPFC) (MNI: x = 8, y = 38, z = −2) during trust choices (Fig. 5B) and precuneus after a trust choice (Fig. 5C) were associated with individual τ parameters. Further, willingness to trust predicted model-based expectations (ρ_*t*_) related ACC activity at the beginning of the trial (Fig. 5D) and activity in the caudate (extending to putamen), posterior cingulate gyrus and angular gyrus modulated by model-based action utilities during trust choices (Fig. 5F **& Tab. S1**). In contrast, willingness to trust was negatively correlated with VMPFC activity associated with trialwise trust choices (Fig. 5E) but positively with right AI activity associated with average payoffs during feedback (Fig. 5G **& Tab. S2**). These results suggest that brain regions more engaged in controls than SZ patients (e.g., putamen, caudate, and AI) were also modulated by an individual’s willingness to trust. Importantly, brain activity in the right putamen was more strongly associated with willingness to trust in controls than individuals with SZ (Fig. 6A) and similar results were found for brain activity in the precuneus and bilateral putamen associated with model-based action utilities (Fig. 6B).

**Figure 5.**
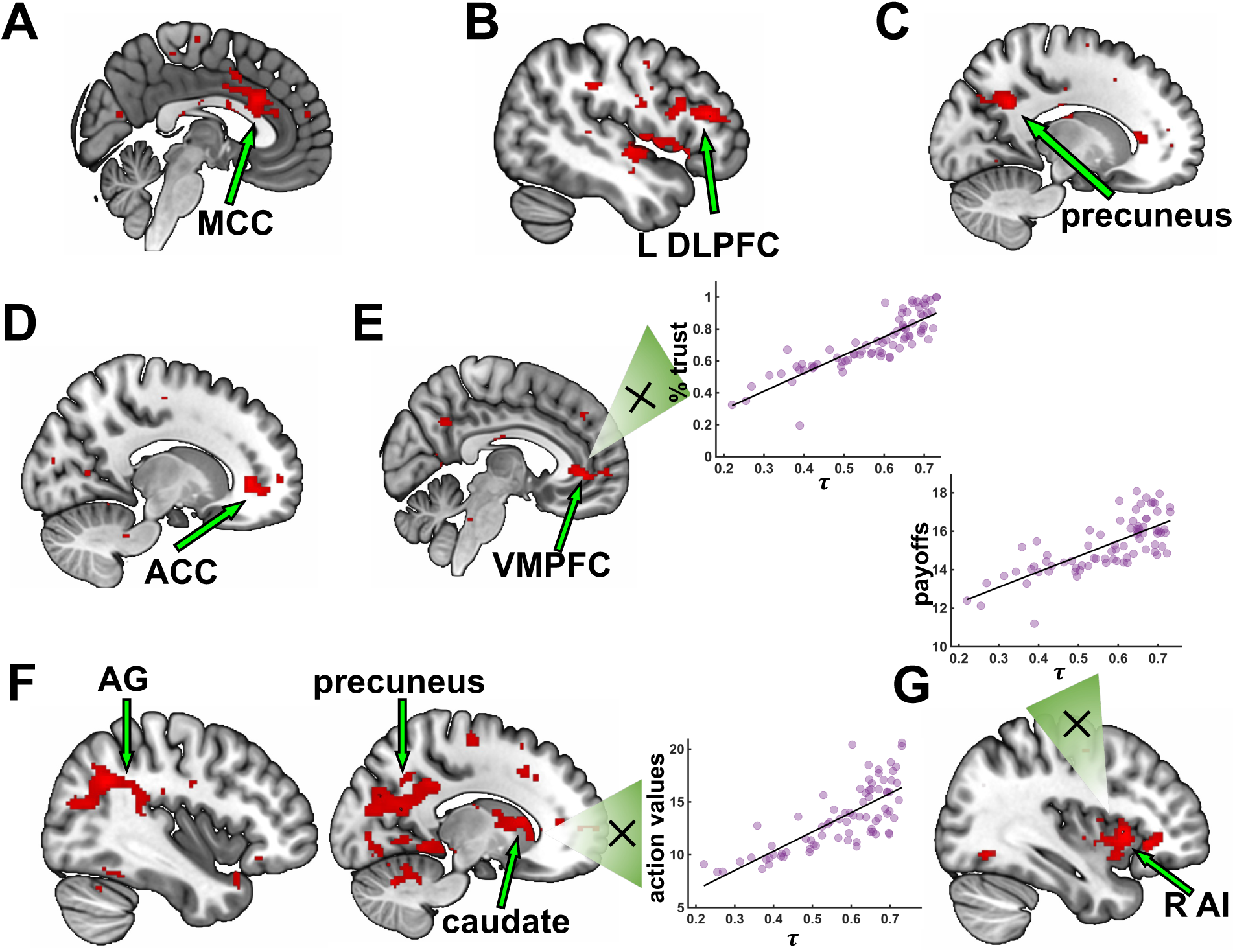
Willingness to trust modulates brain activity. **A.** Willingness to trust (τ) modulated MCC activity (MNI: x = 2, y = 22, z = 26) at investment cue (beginning of the trial). **B.** Activity in the left DLPFC (MNI: x = −46, y = 32, z = 14) during trust choices was positively modulated by willingness to trust. **C.** At trust, willingness to trust modulated activity in the precuneus (MNI: x = −14, y = −54, z = 30). **D.** ACC activity (MNI: x = 14, y = 38, z = 2) associated with individual expectations (ρ_*t*_) at the beginning of the trial was modulated by willingness to trust (τ). **E.** VMPFC activity (MNI: x = 8, y = 38, z = −2) associated with investments during trust choices was negatively modulated by willingness to trust (τ). **F.** Caudate (MNI: x = −6, y = 10, z = 6) and AG (MNI: x = 40, y = −54, z = 30) activity associated with model-based action values during trust choices was positively modulated by willingness to trust. **G.** At feedback, AI activity (MNI: x = 44, y = 16, z = −4) associated with own average payoffs was positively modulated by willingness to trust. MCC, middle cingulate cortex; DLPFC, dorsolateral prefrontal cortex; ACC, anterior cingulate cortex; VMPFC, ventromedial prefrontal cortex; AG, angular gyrus; AI, anterior insula; L, left; R, right. FWE<.05 cluster-level corrected.

**Figure 6.**
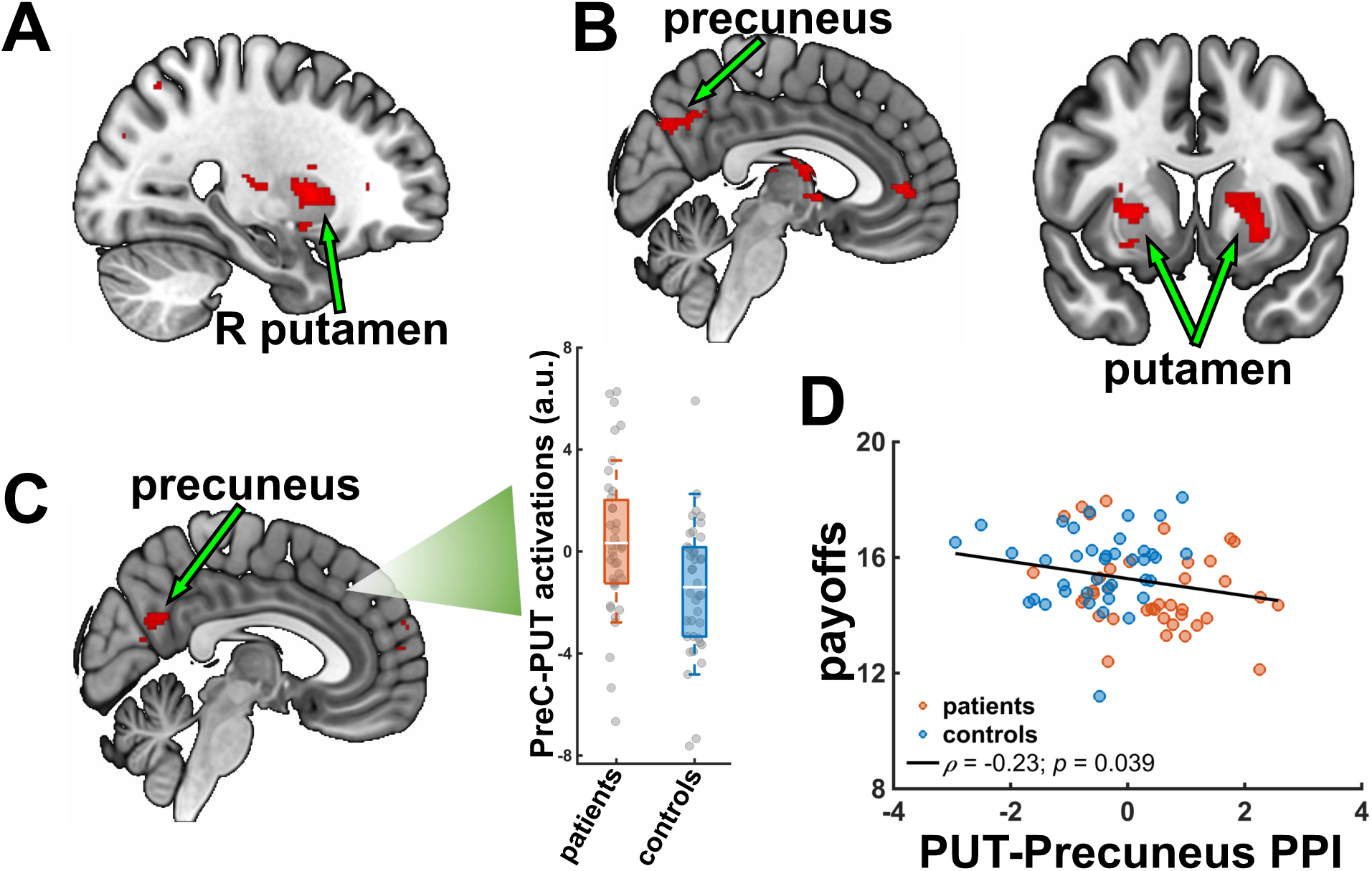
Functional connectivity results. **A.** During trust choices, the right putamen (MNI: x = 26, y = 6, z = 4) was more strongly modulated by willingness to trust in controls than SZ patients. **B.** Activity in precuneus (MNI: x = 18, y = −54, z = 34), right (MNI: x = 26, y = 6, z = 2) and left (MNI: x = −24, y = 10, z = −4) putamen associated with model-based action values was more strongly modulated by willingness to trust in controls than SZ patients. **C.** Right putamen manifested stronger functional connectivity with precuneus (MNI: x = 4, y = −58, z = 26) in controls than patients. **D.** Lower functional connectivity between these two brain regions was associated with lower average payoffs in the task. PUT, putamen; PPI, psychophysiological interaction; R, right.

These results suggest a central role for the putamen in evaluations of a decision to trust, the weighting of an individual’s expectations of trustee reciprocity, and learning to trust from feedback about the benefits of trust. To gain further insights into the function of this brain region, we investigated its functional pathways with the rest of the brain using functional connectivity analyses. A whole-brain psychophysiological interaction revealed that the right putamen was more strongly connected with the precuneus in controls than individuals with SZ during feedback (Fig. 6C). In particular, lower activity in the putamen was compensated by stronger engagement of the precuneus in individuals with SZ (Fig. 6C, insert), leading to a stronger negative functional connectivity in individuals with SZ as compared to controls. Further, this negative functional connectivity negatively correlated with individual payoffs across groups (*r*_s_ = −0.23, *p* = .039; Fig. 6D). Given the above associations of the precuneus with action utilities and willingness to trust during trust choices, this functional connectivity between putamen and precuneus suggests these brain regions are pivotal to enabling trust and the establishment of successfully trusting relationships by enabling accurate integration of information for belief updating in social interactions.

## Discussion

In this study, we investigated trust dynamics during interactions with benevolent partners in a group of individuals with SZ and healthy controls. We observed that individuals with SZ exhibited greater distrust in benevolent partners than controls. This misplaced distrust by individuals with SZ was due to their failure to adequately adjust their behavior to trustee reciprocity. Computational modeling showed that individuals with SZ were less inclined to rely on expectations about trustee reciprocity, which was associated with reduced activity in the putamen and precuneus. Furthermore, stronger functional connectivity between these regions was linked to less successful trusting interactions in individuals with SZ with overall lower payoffs, suggesting that this neural circuit plays a key role in learning to trust in individuals with SZ.

Consistent with prior research on psychosis^10^, individuals with SZ were on average more distrustful than controls. However, initial trust levels did not differ between the groups. Instead, reduced sensitivity to changes in reciprocity led to flat trust levels throughout the interaction in individuals with SZ. In contrast, controls adequately reacted to trustee reciprocity and gradually increased their trust over time. Notably, healthy participants were not merely reacting to the partner’s behavior, but they also appeared to strategically encourage reciprocity, as seen in stronger trust increases following decreasing reciprocity levels (known as *coaxing*). In contrast, distrustful individuals with SZ were less likely to increase their trust in response to decreasing reciprocity, and if they did make themselves more vulnerable by investing more, they were more likely to sharply reduce their trust afterward, leading to highly variable investment behaviors. Although choice variability has received little attention in mental health research, some studies link it to impaired decision-making processes. For example, increased decision noise and choice-switching has been found in individuals with SZ during information-sampling and reward-learning tasks^31–33^. Further, variability in metacognitive decisions has been associated with abnormal confidence—a known feature of schizophrenia and other mental disorders^34^––and with compulsivity and intrusive thoughts as measured by questionnaires^35^.

Together, these results point to two mechanisms contributing to reduced trust in individuals with SZ. First, heightened caution in their decisions prevented them from sufficiently probing their partner’s trustworthiness and gradually building trust. This aligns with findings showing lower trait-like interpersonal trust in individuals with SZ (as measured by a questionnaire)^36^. Second, individuals with SZ showed greater discomfort with vulnerability (i.e., high investments) and tended to sharply reduce their trust in future interactions to avoid the risk of betrayal. This pattern mirrors findings in disorders such as borderline personality disorder, where sustaining cooperation in trusting relationships is also impaired^20^. Finally, choice variability stood out as a potentially domain-general signature of cognitive impairments impacting different behaviors in social and non-social contexts.

Computational modeling results provided further insights into these model-agnostic results. Specifically, participants who exhibited greater trial-by-trial choice variability were less likely to base their trust on expectations of trustee reciprocity. In individuals with SZ, this was particularly pronounced: not only were they generally less willing to trust, but their reduced willingness to trust (model-based τ parameter) was also associated with greater trialwise behavioral sensitivity to both positive and negative reciprocity. This pattern suggests that investment behavior in individuals with SZ was more heavily influenced by the fear of a potential betrayal than by informed beliefs about the trustee’s benevolence learned from the history of the social interaction. Indeed, model-based action utilities were also consistently lower in SZ patients, suggesting that the negative consequences of trust loomed larger on their investment decisions and impeded their ability to increase their levels of trust. These findings align with previous evidence on impairments in complex social inference in SZ^37^ and help explain why patients failed to adapt to the consistently benevolent behavior of our preprogrammed trustee. Rather than relying on their beliefs about their partner’s reciprocity based on evidence from past behavior, individuals with SZ disproportionately feared being vulnerable to the trustee by putting more weight on the potential of a betrayal, thereby undermining the possibility of forming a sustained trusting relationship.

Moreover, different behavioral strategies may underlie SZ patients’ greater choice variability. One possibility is that patients adopted a simpler “all-or-nothing” heuristic than other more computationally-intensive inference processes, choosing to either manifest high levels of trust or radical distrust. Alternatively, their decisions may have been characterized by general inconsistency or noise. Group comparisons of choice stochasticity parameters ( χ policy parameter) revealed that SZ patients exhibited higher decision noise, which impaired their ability to elicit greater reciprocity from the trustees, reducing the effectiveness of their social learning. This resembles previous findings from reversal learning tasks, where SZ patients demonstrate a higher tendency to switch choices, leading to poorer learning outcomes^38^.

Importantly, though, these noisier choice patterns could also be driven by SZ individuals’ tendency to “jump to conclusions”^39^. This could in turn be due to their perception that the environment is highly volatile, which can impact choice values uniforming the value distribution over available options^32^. When the values of different choice options become more similar, the resulting behavior appears more random. This is the effect induced in our study by reduced willingness to trust in individuals with SZ, which contributed to lower action utilities and reduced expected-value differences between investment choices in individuals with SZ. These effects can lead to noisier and suboptimal choice behaviors, previously associated with poorer learning performances^40,41^. Hence, impaired learning in SZ is likely to arise from suboptimal, noisy choice strategies leading to maladaptive information sampling from the social environment that hinders the acquisition of stable, reliable beliefs about others^42,43^.

Reduced willingness to trust as estimated by our computational model is akin to underweighting expectations of trustee reciprocity—consistent with a diminished influence of prior beliefs on current decisions. This pattern parallels the effects of reduced prior precision in Bayesian inference, a core predictive processing abnormality proposed in psychosis^44^. In this framework, SZ individuals’ choices appear less influenced by the partner’s distant, past behavior, as would have been expected if priors about trustee reciprocity had been more robustly weighted. This interpretation aligns with our model-agnostic findings indicating a reduced choice-history bias in individuals with SZ and with previous work demonstrating similar effects in tasks involving serial presentations of non-social stimuli^45^. Importantly, behavioral inconsistency may also contribute to this underweighting of priors in social contexts. Specifically, high variability in choice behavior introduces greater noise in the empirical observations (likelihood). As a result, individuals with SZ may accumulate evidence of lower signal quality across time. This did not happen in our task, as we preprogrammed our trustees to be stubbornly benevolent. Future studies could test whether in social interactions, variable behavior in individuals with SZ might confuse their interacting partners, making them manifest contradicting responses. In sequential Bayesian inference, integration of noisy evidence leads to more uncertain beliefs that amount to reduced prior precision over time^46^, which could explain the reduced contribution of expectations of trustee reciprocity to trust decisions in our SZ individuals.

Importantly, SZ individuals’ difficulty in overcoming distrust was reflected in heightened engagement of the AI and weaker recruitment of the striatum during the trust phase. The AI has commonly been implicated in psychopathology^47,48^ and its functional role spans from risk perception and loss expectations in non-social tasks^49,50^ to fear of betrayal during social interactions with unfamiliar partners^51^. In the investment game, AI activation has been associated with concerns about future resource distribution inequalities—both disadvantageous (e.g., regret) and advantageous (e.g., guilt)^18,51–53^. Notably, increased AI activity serves as a cautionary signal in early stages of trust development but typically diminishes as individuals get to know their partner’s behavior and begin to rely more on striatal engagement^54,55^. In contrast, previous studies have consistently shown that trust elicits stronger neural activations in reward-processing brain regions like the striatum and that these brain regions track individual levels of trust^20^. Hence, lower engagement of the striatum in our individuals with SZ could underpin a failure to anticipate the positive consequences of trust, leading to lower trust levels. On the contrary, sustained hyperactivity in the AI in our individuals with SZ could relate to higher uncertainty about the partner’s trustworthiness and, hence, heightened sensitivity to the potential negative consequences of trust, impairing accurate trustworthiness learning.

Parametric modulation analyses further support this interpretation. In controls, activity in the striatum (caudate and putamen) and precuneus correlated more strongly with model-based action utilities, and this neural pattern was positively associated with an individual’s willingness to trust. These brain regions––known to support social learning and prediction––have been found to show altered activation and connectivity in first episode and chronic SZ^56–58^. For example, the precuneus is less responsive to outcome uncertainty during nonsocial learning in individuals with SZ compared to controls^59^. Consistent with this finding, in our study, controls exhibited stronger precuneus engagement, and this activity positively correlated with greater willingness to trust, indicating that the precuneus is central to anchoring social behaviors to inferences about others. Moreover, in our study, the putamen appears to play a critical role in varying cognitive processes, such as the evaluations of trust decisions, the weighting of expectations about others’ trustworthiness, and feedback learning. Crucially, this brain region manifested stronger negative functional connectivity with the precuneus in individuals with SZ compared to controls. Previous studies have shown that cortico-striatal dysconnectivity in SZ is associated with deficits in learning, cognitive performance, and symptom severity^60–62^, and, in particular, altered functional connectivity between the precuneus and the striatum correlates with clinical SZ symptoms^63^. In addition, the negative functional connectivity between the putamen and the precuneus in our study was further linked to lower payoffs, indicating a pivotal role in effective social learning during trusting interactions. The interplay between these two brain regions might represent a compensatory dynamic, whereby suboptimal feedback learning (via reduced putamen activity) could be offset by stronger engagement of inference processes (via increased precuneus involvement). Importantly, while increased inference processes could compensate for impaired trial-by-trial learning, this compensatory mechanism may further promote the paranoid thinking patterns characteristic of SZ^64^.

Taken together, we showed the multifaceted factors underlying misplaced distrust in a sample of SZ individuals and healthy controls. We found that lower trust in SZ individuals is based on an underweighting of beliefs about their partner’s benevolence, whereby the prospects of negative outcomes weighed more heavily on their investment choices than their partner’s benevolent reputation. Greater behavioral variability in individuals with SZ might lead to variable social feedback that impairs interpersonal learning and, in turn, hinders the formation of reliable expectations necessary for establishing trust. Heightened recruitment of AI and weaker engagement of the striatum in SZ individuals might reflect their tendency to underweight partner benevolence and contribute to increased sensitivity to the potentially negative outcomes of trust decisions. Taken together, these findings shed light on the cognitive and neural mechanisms that give rise to distrust in SZ, offering a more nuanced understanding of the processes that shape social decision-making in individuals with this disorder.

## Materials and Methods

### Participants

This study recruited 39 individuals with a diagnosis of schizophrenia (according to ICD-10 criteria) (22-60 years; *M* = 42.92, *SD* = 10.0) and 39 healthy controls (23-55 years; *M* = 34.05, *SD* = 9.76) matched for age, sex, and socioeconomic background. All patients were currently receiving antipsychotic medication, individuals prescribed clozapine were excluded. Exclusion criteria for all participants were a history of neurological illness, current major physical illness, and drug dependency over the last six months. Exclusion criteria for controls were a history of psychiatric illness and a first-degree relative having suffered from a psychotic illness. All subjects had normal hearing and normal or corrected-to-normal vision. The study was approved by the London Camberwell St Giles Research and Ethics Committee and all participants provided written informed consent before the experiment.

### Task

To measure trust, a multi-round investment game was used (Fig. 1A)^65^. In this neuro-economic computer-based game, consisting of 20 trials, participants played the role of the investor. At the start of each trust game trial the initials of the other player were displayed on the computer screen. Participants (investors) received an initial endowment of £10 and indicated, on a horizontal scale from zero to ten pounds, how much they wanted to share with the other player (trust). The chosen amount was tripled during the transaction and after this the trustee made a repayment (reciprocity). At the end of the trial, they saw the kept and given amounts and the total earnings for both players. After this a new trial started.

The computer-simulated trustee was pre-programmed to behave in a probabilistically benevolent manner. In particular, on each first repayment, the return could be 1.0, 1.5, or 2.0 times the initial investment, with each outcome equally likely (i.e., 33%). Starting from the second trial, the probabilities of these repayment factors were adjusted based on changes in the investor’s behavior—specifically, whether they increased their investment compared to the previous round. If the investor raised their investment or consistently invested the maximum amount (£10), the probability of receiving the highest return (2.0) increased by 10%, the probability of the lowest return (1.0) decreased by 10%, and the probability of the middle return (1.5) remained unchanged. This adjustment continued across trials until the probability of the highest return reached a maximum of 63%. This algorithm simulated the behavior of a benevolent trustee by rewarding increases in trust with greater reciprocity, while decreases in trust had no negative impact on repayments.

### Model-agnostic analyses

Mixed-effect Bayesian regressions were implemented in model-agnostic analyses. One regression predicted trialwise trust (investments) using reciprocity on the previous trial, group, and time (trial number) as fixed-effects regressors with a random slope for participants. A similar regression was implemented to predict trust changes as the difference in trust between the current and the previous trial. Previous trust, group, and the difference between reciprocity in the last trial and two trials back were used as regressors. To test for time effects, a time regressor was further introduced coding trial number, allowing for a random intercept and slope for participants. A further regression to check our algorithm’s behavior was run with the difference between the previous and current reciprocity predicting participant’s last trust change. For all models, we used uninformative, Gaussian beta priors, i.e., β∼𝒩(0,5). Stan was used to fit models with the no-U-turn sampler for efficient exploration of posterior estimates (https://mc-stan.org) with four chains, 1000 tuning steps burn-in steps and 6,000 samples. The 95% highest posterior density intervals were calculated from these samples. Visual inspection of the traces and the Gelman-Rubin statistics (*R*^K^) were used to assess convergence^66^.

### Computational modeling analyses

We used Approximate Bayesian Computation (ABC) to hierarchically fit our computational model employing population Monte Carlo sampling^67^. In ABC, we first sample candidate parameter values for group-level prior distributions from which subject-specific parameters were sampled. For the winning model, beta distributions were used to estimate prior expectations and τ parameters (which were defined to be 0 ≤ τ ≤ 1) and a normal distribution for the inverse temperature. We then use these candidate parameters to generate 100 simulations of participants’ choices in the task. These simulations are then compared to the observed data using root-mean-squared deviation as a distance function (negative log-likelihood). If the distance measure was smaller than a pre-set error threshold ε_*t*_, we accepted the candidate sample, otherwise we rejected it. We used a set of exponentially decreasing error thresholds ε_*t*_ with values *e*^(1.5:−.1:1.5)^ . We ran 100 permutations with 1000 particles each. Model fit of the different computational models was compared using Bayesian Information Criterion (BIC) that allows us to balance model performance against model complexity. Simulated behavior based on fitted parameters strongly correlated with actual behavior for both individuals with SZ (ρ_37_ = .91) and healthy controls (ρ_37_ = 96). Parameter recovery shows very good recoverability for the τ parameters with average correlations between estimated and recovered values of *r* = .45 for both τ_*p*_ and τ_*n*_ (all *p*s < .0005). Average correlation values for the prior (*r* = .34, *p* < .007) and the inverse temperature (*r* = .45, *p* < .001) were similar and also highly recoverable (all *p*s < .0005).

Finally, to test for group differences in model parameters, a participant-level Bayesian regression was run with the same uninformative priors as above and using group and parameter type (i.e., τ_*p*_ or τ_*n*_) as predictors for τ parameter values.

### Neuroimaging analyses

#### Scanning parameters

Functional images were collected using a T2*-weighted echo planar imaging (EPI) sequence optimized for blood oxygenation level-dependent (BOLD) contrast (430 volumes; repetition time [TR] = 2000 ms; echo time [TE] = 35 ms; field of view = 240 mm; slice thickness = 3 mm; matrix size = 64 × 64; flip angle = 75°) on a 3T GE Excite II MRI system (GE Healthcare, USA). High-resolution structural images were also acquired for each participant using a T1-weighted magnetization-prepared rapid gradient echo (MP-RAGE) sequence (TR = 7321 ms; TE = 3 ms; inversion time [TI] = 400 ms; field of view = 240 mm; slice thickness = 1.2 mm; 196 slices).

#### Image preprocessing

Neuroimaging data were preprocessed using a standard pipeline in SPM12 (v. 6905; http://www.fil.ion.ucl.ac.uk/spm/software/spm12/) in MATLAB R2022b (The Mathworks, Natick, Massachusetts; http://www.mathworks.com/). The functional images were corrected for slice acquisition time, realigned for head movement correction to the mean image, co-registered to their structural images using the unified segmentation procedure^68^, normalized into MNI space using deformation fields from the segmentation procedure (resampling voxel size: 2×2×2 mm^3^), and spatially smoothed using a Gaussian filter (8×8×8 mm^3^ full width at half maximum, FWHM) to decrease spatial noise.

#### First-level neuroimaging analyses

At the first level, a general linear model (GLM1) was estimated with five regressors to test for main effects of each of the task phases (i.e., investment cue, trust, trust choice, reciprocity, feedback; see Fig. 1B). GLM2 further contained parametric modulators with behavioral and model-based parameters. For the trust choice and reciprocity phases, GLM2 had amounts invested (trust) and returned (reciprocity) as parametric modulators, respectively. For the feedback phase, GLM2 had amounts earned in that trial by the two partners as parametric modulators. With respect to model-based parameters, GLM2 had trialwise expectations as parametric modulator of the investment cue phase, trialwise action utilities as parametric modulator of the trust choice phase, and expectation violation (i.e., the difference between model-based expectations of trustee reciprocity and observed current reciprocity) and surprise (i.e., negative log likelihood of individual beliefs about trustee reciprocity on a trial-by-trial basis) as parametric modulators of the reciprocity phase. Moreover, motion parameters were included as regressors of no-interest. A temporal high-pass filter with a cutoff of 128 s was applied. Contrast analyses between beta regressors were performed on the first level and later used for second-level whole-brain analyses.

#### Second-level neuroimaging analyses

To examine neural signatures of trusting behaviors and trustee reciprocity learning, one-sample *t*-tests on main effects and parametric modulators were performed at the second (group) level. To study group differences, contrast images from the first level were entered into a two-sample *t*-test. Results were whole-brain corrected for multiple comparisons using a voxel-defining threshold of *p* < .005 and a family-wise error of *p* < .05 (cluster-level corrected)^69^.

#### Whole-brain regression analyses

To investigate the relationships of individual willingness to trust (τ parameter estimated for each participant by the winning computational model) with neural responses in the game, contrast images from the first level were regressed against τ parameter values in a group-level whole-brain regression analysis (using the same multiple comparison parameters as above).

#### Task-dependent functional connectivity analysis

To investigate the potential functional pathways of the putamen, a task-dependent functional connectivity analysis was implemented using a whole-brain psychophysiological interaction analysis (PPI)^70,71^ with the significant putamen cluster derived from the main effect of the trust choice phase as seed region (10 mm radius). The PPI-GLM consisted of a task regressor, a physiological regressor entailing deconvolved blood-oxygen-level-dependent (BOLD) signal from the seed region and a regressor for the interaction term. Movement parameters were entered as regressors of no interest. Significant connectivity was assessed with the same multiple comparison parameters as above.

#### Labeling and data visualization

Anatomical labeling was based on the SPM Anatomy toolbox (v. 2.2)^72^ and MRIcron (http://people.cas.sc.edu/rorden/mricron/install.html/). Visualizations were done using MRIcroGL (https://www.mccauslandcenter.sc.edu/mricrogl/home/).

## Data Availability

The behavioral data and code are openly available on OSF (https://osf.io/gfspd/overview?view_only=a4559ce856b343049aa03c1c60b2bd17).

## Acknowledgments

SSS is funded by Medical Research Council grant (MR/V037218/1) and a Wellcome Trust mental health award and some of this work was funded by an European Research Council Consolidator award (MUTRIPS no.311686). We would like to acknowledge the support of Szentgyorgyi T, Crisp C, Mouchlianitis E in the original MUTRIPs study.

